# Effect of treatment with GLP-1R agonists on the urinary peptidome of T2DM patients

**DOI:** 10.1101/2023.05.03.23289439

**Authors:** Sonnal Lohia, Justyna Siwy, Emmanouil Mavrogeorgis, Susanne Eder, Stefanie Thoeni, Gert Mayer, Harald Mischak, Antonia Vlahou, Vera Jankowski

## Abstract

**Aim:** Type II Diabetes mellitus (T2DM) accounts for approximately 90% of all DM cases in the world; and is diagnosed with increased levels of blood glucose or hyperglycemia. Glucagon-like peptide-1 receptor (GLP-1R) agonists have established an increased capability to target directly or indirectly six core defects associated with T2DM, while, the underlying molecular mechanisms of these pharmacological effects are not fully known. This exploratory study was conducted to analyze the effect of treatment with GLP-1R agonists on the urinary peptidome of T2DM patients.

**Material and Methods:** Urine samples of thirty-two T2DM patients from the PROVALID study (A Prospective Cohort Study in Patients with T2DM for Validation of Biomarkers) were collected at pre- and post-treatment with GLP-1R agonist drugs, and analyzed by capillary electrophoresis coupled to mass spectrometry (CE-MS).

**Results:** In total, 329 urinary peptides were identified, of which 70 were significantly affected by the GLP-1R agonist treatment; fragmenting from 26 different proteins. The downregulation of MMP proteases, based on the concordant downregulation of urinary collagen peptides with GLP-1R agonist treatment was highlighted in the study. Treatment also resulted in the downregulation of peptides from SERPINA1, APOC3, CD99, CPSF6, CRNN, SERPINA6, HBA2, MB, VGF, PIGR and TTR, many of which were previously found associated with increased kidney and/or vascular damage, indicating beneficial effect of GLP-1R agonists.

**Conclusions:** The novel findings in this study, indicate potential molecular mechanisms of GLP-1R agonists in the context of management of T2DM and prevention or delaying the progression of its associated diseases.

## Introduction

In 2021, the International Diabetes Federation (IDF) predicted a five-fold increase in the worldwide adult population of Diabetes mellitus, from 150 million in 2000^1^ to 783 million in 2045^2^; of which Type II Diabetes mellitus (T2DM) is said to account for approximately 90% of all cases^3^. T2DM is characterized as a chronic disease^4^ and is diagnosed with increased levels of blood glucose or hyperglycemia^5^. T2DM has been associated with macrovascular complications, such as atherosclerosis, myocardial infarction and diabetic foot syndrome; as well as microvascular complications, such as neuropathy, retinopathy, and nephropathy^6-8^. Approximately 44% of all T2DM cases have been linked to obesity, correlating an exponential increase in their worldwide prevalence^9,10^. Additionally, the World Health Organization (WHO) predicts 57.8% of the world population to be overweight and obese by 2030^11^.

Since no cure exists, efforts to control and treat T2DM have escalated^12^. The American Diabetes Association (ADA) recommends healthy lifestyle changes as the first-in-line response, followed by metformin-based pharmaceutical interventions^13^. The approach of lowering glucose plasma levels, remains the most sought-after treatment and has recently witnessed novel and effective advances. These alongside metformin, commonly include treatment with glucagon-like peptide-1 receptor (GLP-1R) agonists, sodium-glucose co-transporter-2 (SGLT-2) inhibitors, dipeptidyl peptidase-4 (DPP-4) inhibitors and insulin pumps followed by continuous monitoring of glucose levels^8^. Furthermore, research focusing on treatment paradigms targeting the ‘ominous octet’ of T2DM, implicated in the development and progression of hyperglycemia, has gained momentum in the last couple of decades.

The GLP-1R agonists have dramatically transformed patient care guidelines for T2DM^14^. In comparison to the other antihyperglycemic medications, GLP-1R agonists have established an increased capability to target directly or indirectly six out of the eight core defects associated with T2DM^15-17^. In response to food intake, the intestinal L-cells dependent secretion of GLP-1 is significantly impaired in T2DM patients, which affects the pancreatic β cells dependent insulin secretion; eventually resulting in hyperglycemia^18-20^. The synthetically produced GLP-1R agonists stimulate GLP-1R, in a similar fashion as the native GLP-1. Treatment benefits of peptide-based GLP-1R agonists include glycemic control, weight loss, delayed onset of macroalbuminuria, prevention of clinical hyperglycemic episodes and cardiovascular events, and reduced estimated glomerular filtration rate (eGFR) decline; that in turn indirectly result in the inhibition of glucagon secretion and increased secretion of insulin with minimal hypoglycemic risks^8,21-23.^

Since their first approval in 2005, seven GLP-1R agonists drugs have been developed for glycemic control in T2DM patients. In 2021, Trujillo *et al*., described fourteen head-to-head trials conducted on these seven GLP-1R agonists, the results of which continue to establish the benefits of this class of drugs in treatment of T2DM patients. The authors further highlighted studies that have published beneficial cardiovascular and renal outcomes of the treatment; however, the results, in terms of the magnitude for glycemic control and adverse effects are not consistent^24^. The focus of research thus needs to shift from investigating the pharmacological effects of GLP-1R agonists, to understanding the underlying molecular mechanisms. Therefore, in this exploratory study, we aimed to assess the effect of treatment with GLP-1R agonists on the urinary peptidome of T2DM patients, in an untargeted peptidomics approach.

Recent advances in the analyses of urinary proteome and peptidome, have paved way for the detection of alterations at both pathological as well as physiological levels in chronic diseases^25^. Urine-based omics studies have especially garnered relevance, due to the ease of sample collection, longer stability of the peptides, complex composition (from blood, kidney, and bladder) and the possibility of larger cohort studies because of its non-invasive approach. In addition, the water soluble and charged nature of urinary peptides enable their uncomplicated mass spectrometric (MS) based detections^26^. The urinary peptidomic analysis in this study was performed by a capillary electrophoresis coupled to mass spectrometry (CE-MS) technique, which supports the identification of naturally occurring urinary peptides and peptidomic changes in response to drug interventions^27^.

## Materials and Methods

### Study population and sample collection

Used were urine samples of thirty-two T2DM patients, administered renin-angiotensin system (RAS) inhibitor medications at the primary health care level, recruited within the Prospective Cohort Study in Patients with Type 2 Diabetes Mellitus for Validation of Biomarkers (PROVALID) study. PROVALID is an observational, prospective cohort study in five European countries, as described previously^28^. Consent was obtained from all the patients. Urine samples were collected from T2DM patients at their visit just before GLP-1R agonists prescription (Liraglutide 1.2-6 mg/ml or Exenatide 0.01-0.25 mg/ml or Lixisenatide 0.01-0.02 mg/ml) and labelled as pre-treatment samples. The urine samples collected at the first visit after the treatment initiation were labelled as post-treatment samples. All urine samples were stored at -20°C until analysis.

### Sample preparation and CE-MS analysis

The standard operating protocols describing urine sample preparation and extraction of peptides followed in this study, have been previously published^29^. The CE-MS analysis was conducted as described in detail by Zurbig *et al*.,^30^ utilizing a P/ACE MDQ capillary electrophoresis system (Beckman Coulter, Fullerton, CA, USA) coupled with a Micro-TOF II MS (Bruker Daltonic, Bremen, Germany) instrument. Literary evidence on the advantages of CE-MS analysis in terms of reproducibility, sensitivity, precision and accuracy are extensively available^31^. The relative peptide intensities were normalized based on an internal standard of 29 stable collagen peptides, that can be detected naturally in urine samples of healthy and diseased. This calibration was performed for normalizing the variability in peptide intensities^32^. The resulting peptides and their normalized intensity values were stored in an internal Microsoft SQL database^33^, which enabled the comparison of the pre- and post-treatment urinary peptide profiles. For identification of the peptide sequences, MS/MS based analysis by a Dionex Ultimate 3000 RSLS nanoflow system (Dionex, Camberley, UK) or a Beckman P/ACE MDQ CE that was coupled to an Orbitrap Velos MS instrument (Thermo Fisher Scientific Inc., Boston, MA, USA) was performed, as described previously^34^.

### Statistical analysis

All statistical analyses in this study were performed using R programming (R version 4.2.2, R Foundation for Statistical Computing, Vienna, Austria with IDE: R Studio Version 1.2.5, Boston, MA, USA). As a pre-requisite for the analysis, thresholds of 30% peptide frequency (in at least one treatment group) and area under the curve (AUC) value ≥ 0.60 were applied. The normally distributed and continuous datasets generated from CE-MS based peptide profiles of the urine samples, obtained from pre-treatment (*n*=32) and post-treatment (*n*=32); were compared by a paired Wilcoxon rank-sum test, using the row_wilcoxon_paired() function from the matrixTests package. A *p*-value < 0.05 was considered statistically significant, which was further adjusted for false discovery rates (FDR) by the Benjamini-Hochberg method^35^. All the plots in this manuscript were created using the ggplot() function from the ggplot2 package.

### Bioinformatic - Proteasix analysis

The proteolytic events responsible for the secretion of the statistically significant urinary peptides were investigated by the open-source online tool “Proteasix” (http://www.proteasix.org, accessed on 6 March 2023). In brief, Proteasix retrieves information from literature and databases like MEROPs, UniProt Knoweldgebase (KB) and CutDB. The “Observed Prediction tool” of Proteasix with search parameters set to default, was utilized in this study.

## Results

### Study design

The exploratory and untargeted urinary peptidomic approach in this study, was conducted to analyze the effect of GLP-1R agonist treatment on the urinary peptidome of T2DM patients, as illustrated in Figure 1. Briefly, urine samples from thirty-two T2DM patients were collected at two time points: pre-treatment and post-treatment, with a time interval of 11.5 ± 3.72 months in between. Intervention with GLP-1R agonist was introduced at 4.4 ± 4.11 months from pre-treatment visit and the treatment duration until the post-treatment visit was 6.9 ± 3.59 months.

**Figure 1:**
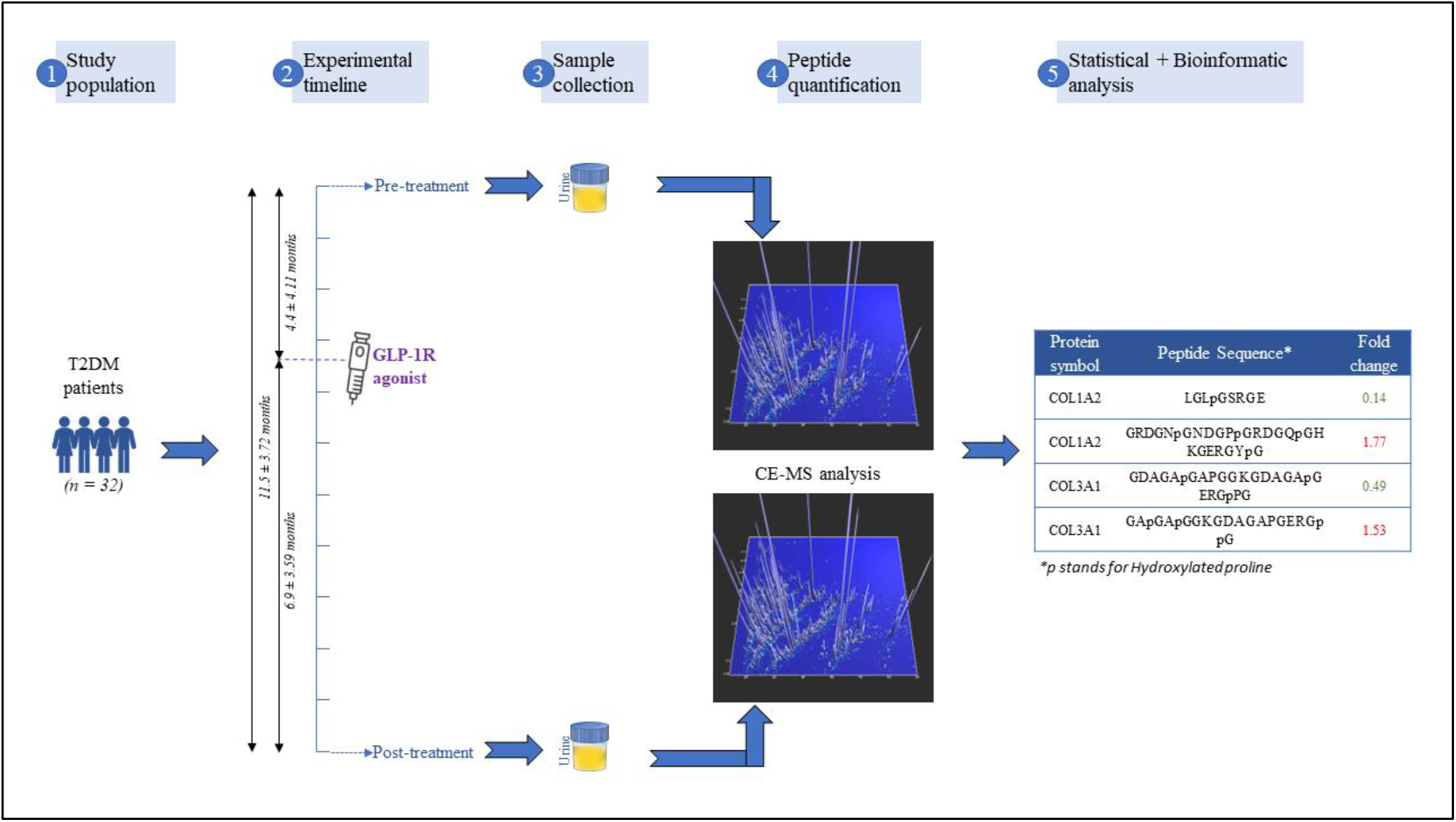
Urine samples from thirty-two T2DM patients were collected at two time points; pre-treatment and post-treatment with the intervention of GLP-1R agonists at 4.4 ± 4.11 months from first sample collection. Naturally occurring urinary peptides were quantified in the urine samples by CE-MS analysis, followed by statistical and bioinformatic analysis of the CE-MS generated urinary peptide profiles.

### Clinical information

The clinical information of all the thirty-two T2DM patients participating in this study, from pre- and post-treatment visits with GLP-1R agonists are provided in the Table 1. The mean age for the patients at first sample collection was 63.7 ± 7.25 years and 56.3% of them were females. As depicted in the Table 1, no statistically significant difference between the clinical levels of Hemoglobin A1C (HbA1C), body weight (BW), body mass index (BMI), systolic and diastolic blood pressure (SBP/DBP), eGFR, albuminuria to creatinine ratio (UACR) and urinary creatinine (UCREA) was observed after the treatment with GLP-1R agonists.

**Table 1:** Clinical information of the T2DM patients in mean ± SD; from pre- and post-treatment with GLP-1R agonists

| *Clinical information* | **T2DM patients (*n*=32)** |  | *p*-value |
| --- | --- | --- | --- |
|  | *Pre-treatment* | *Post-treatment* |  |
| Age (years) | 63.7 ± 7.25 | | |
| Sex (% Females) | 56.3 |  |  |
| HbA1C (%) | 8.0 ± 1.38 | 8.0 ± 1.45 | 0.731 |
| BW (kg) | 98.3 ± 16.13 | 98.8 ± 20.28 | 0.771 |
| BMI (kg/m<sup>2</sup>) | 33.2 ± 6.28 | 33.2 ± 6.93 | 0.915 |
| SBP (mmHg) | 139.2 ± 14.35 | 137.1 ± 12.61 | 0.385 |
| DBP (mmHg) | 81.1 ± 7.74 | 81.3 ± 8.21 | 0.906 |
| eGFR (ml/min/1.73m<sup>2</sup>) | 67.7 ± 14.46 | 67.6 ± 15.55 | 0.965 |
| UACR (mg/g) | 33.6 ± 58.55 | 25.5 ± 44.80 | 0.248 |
| UCREA (mg/dl) | 103.9 ± 41.58 | 96.6 ± 50.72 | 0.429 |

HbA1C: Hemoglobin A1C; BW: Body weight; BMI: Body mass index; SBP: Systolic blood pressure; DBP: Diastolic blood pressure; eGFR: estimated glomerular filtration rate; UACR: Albuminuria to Creatinine ratio; UCREA: Urinary Creatinine

### Peptidomic analysis

Following their collection, the urine samples subjected to CE-MS analysis, generated in total sixty-four urinary peptide profiles (pre-treatment, *n*=32 and post-treatment, *n*=32). The normalized urinary peptide profiles were statistically analyzed by Wilcoxon-rank sum test, pair-wise (pre- and post-treatment per patient). A threshold of AUC values ≥ 0.60 along with a frequency threshold of 30% (i.e., ≥ 10 out of 32 datasets) in at least one treatment group, was applied to the peptide data. This analysis resulted in a list of 329 sequenced peptides to be further investigated. Statistical assessment including Benjamini-Hochberg FDR adjustment yielded a list of 70 statistically significant peptides (adjusted *p*-value < 0.05) affected by the GLP-1R agonist treatment, detailed information of which is provided in Table S1. The distribution of peptide intensity of these 70 statistically significant peptides (red spots) amongst the 329 peptides was also examined (Figure 2a). Assessing the distribution in regulation of all the 329 peptides in response to the treatment (Figure 2b), highlighted the downregulation of urinary peptides in the majority of the statistically significant peptides (66 out of 70 peptides).

**Figure 2:**
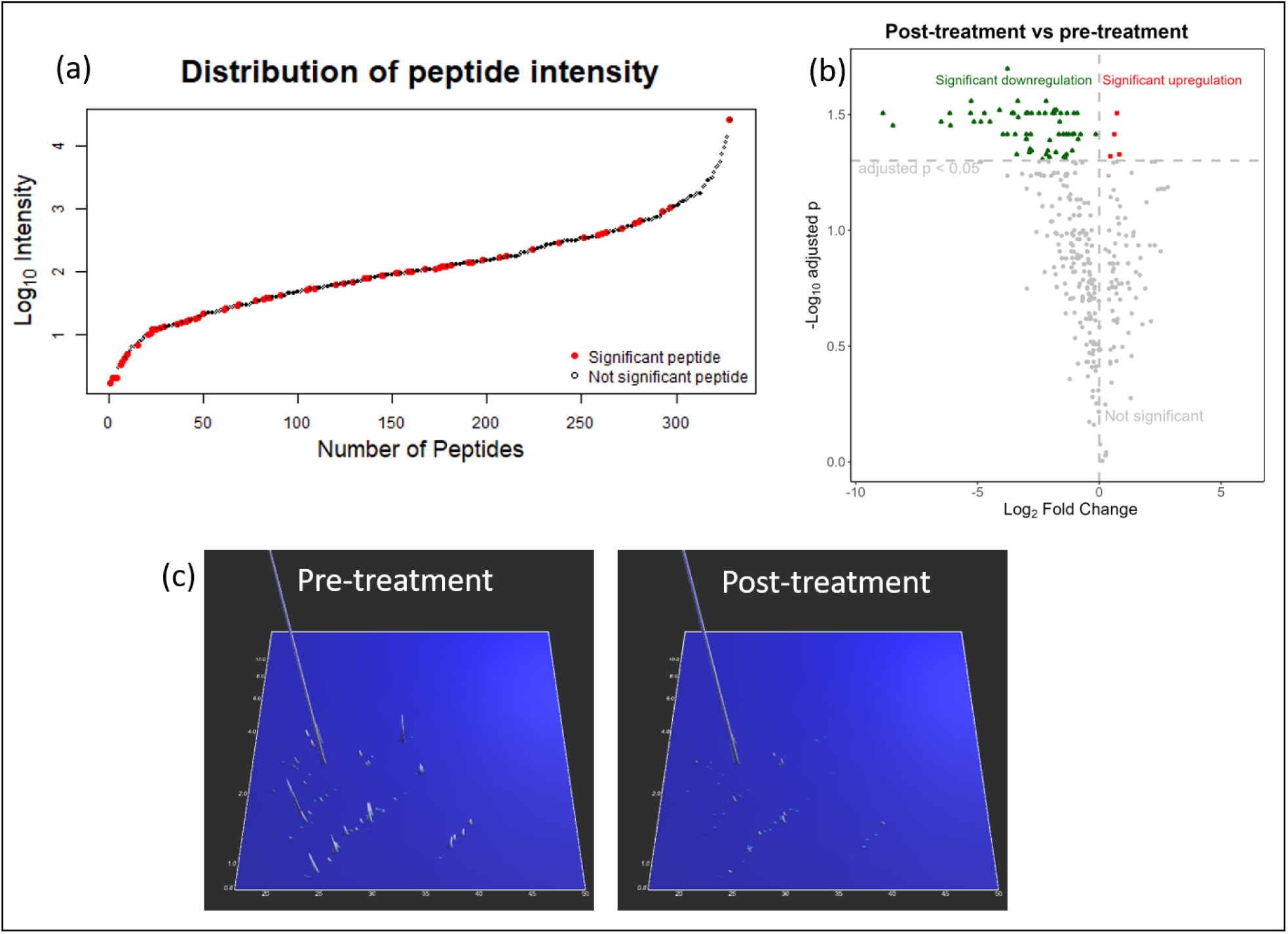
(a) Distribution of peptide intensity for all the 329 sequenced urinary peptides identified in this study. (b) Volcano plot depicting the distribution in regulation of the 329 peptides in response to GLP-1R agonist treatment. (c) The distribution of the 70 statistically significant urinary peptide profiles in pre- and post-treatment of T2DM patients, as obtained from CE-MS analysis.

The 70 statistically significant peptides were identified as fragments of 26 proteins, as shown in Table 2. Most of the peptides (59 out of 70) originated from the collagen family of proteins and 41 out of the 59 collagen peptides belonged in majority to three collagen proteins, namely, collagen alpha-1(III) chain (P02461; COL3A1; *n*=16), collagen alpha-1(I) chain (P02452; COL1A1; *n*=15) and collagen alpha-2(I) chain (P08123; COL1A2; *n*=10). All the remaining identified 11 (out of 70) urinary peptides came from different proteins, alpha-1-antitrypsin (P01009; SERPINA1), apolipoprotein C-III (P02656; APOC3), CD99 antigen (P14209; CD99), cleavage and polyadenylation specificity factor subunit 6 (Q16630; CPSF6), Cornulin (Q9UBG3; CRNN), corticosteroid-binding globulin (P08185; SERPINA6), hemoglobin subunit alpha (P69905; HBA1; HBA2), myoglobin (P02144; MB), neurosecretory protein VGF (O15240; VGF), polymeric immunoglobulin receptor (P01833; PIGR) and transthyretin (P02766; TTR). The 4 out of the 70 urinary peptides showing a statistically significant upregulation on treatment with GLP-1R agonists in T2DM patients, as observed in Figure 2b, were characterized as COL3A1 (*n*=3) and COL1A2 (*n*=1) peptides (Table 2).

**Table 2:** List of 26 proteins yielding the 70 statistically significant urinary peptides, in response to GLP-1R agonists treatment. (↓ refers to down regulation and ↑ refers to upregulation)

| UniProt ID | Gene Symbol | Protein Name | Number of peptides |  |
| --- | --- | --- | --- | --- |
|  |  |  | *Total* | *Regulation after treatment* |
| P02461 | COL3A1 | Collagen alpha-1(III) chain | 16 | ↓ (13) + ↑ (3) |
| P02452 | COL1A1 | Collagen alpha-1(I) chain | 15 | ↓ |
| P08123 | COL1A2 | Collagen alpha-2(I) chain | 10 | ↓ (9) + ↑ (1) |
| P02458 | COL2A1 | Collagen alpha-1(II) chain | 3 | ↓ |
| P02462 | COL4A1 | Collagen alpha-1(IV) chain | 2 | ↓ |
| P05997 | COL5A2 | Collagen alpha-2(V) chain | 2 | ↓ |
| P12107 | COL11A1 | Collagen alpha-1(XI) chain | 2 | ↓ |
| P27658 | COL8A1 | Collagen alpha-1(VIII) chain | 1 | ↓ |
| Q5TAT6 | COL13A1 | Collagen alpha-1(XIII) chain | 1 | ↓ |
| Q05707 | COL14A1 | Collagen alpha-1(XIV) chain | 1 | ↓ |
| Q07092 | COL16A1 | Collagen alpha-1(XVI) chain | 1 | ↓ |
| P39060 | COL18A1 | Collagen alpha-1(XVIII) chain | 1 | ↓ |
| P25067 | COL8A2 | Collagen alpha-2(VIII) chain | 1 | ↓ |
| P29400 | COL4A5 | Collagen alpha-5(IV) chain | 1 | ↓ |
| Q14031 | COL4A6 | Collagen alpha-6(IV) chain | 1 | ↓ |
| P01009 | SERPINA1 | Alpha-1-antitrypsin | 1 | ↓ |
| P02656 | APOC3 | Apolipoprotein C-III | 1 | ↓ |
| P14209 | CD99 | CD99 antigen | 1 | ↓ |
| Q16630 | CPSF6 | Cleavage and polyadenylation specificity factor subunit 6 | 1 | ↓ |
| Q9UBG3 | CRNN | Cornulin | 1 | ↓ |
| P08185 | SERPINA6 | Corticosteroid-binding globulin | 1 | ↓ |
| P69905 | HBA1;<br>HBA2 | Hemoglobin subunit alpha | 1 | ↓ |
| P02144 | MB | Myoglobin | 1 | ↓ |
| O15240 | VGF | Neurosecretory protein VGF | 1 | ↓ |
| P01833 | PIGR | Polymeric immunoglobulin receptor | 1 | ↓ |
| P02766 | TTR | Transthyretin | 1 | ↓ |

Since several peptides from COL1A1 and COL3A1 were found significantly associated with treatment, we investigated the alignment of the peptides in the protein structure. The respective identified peptide sequences were aligned with the primary structure of the proteins, as shown in Figure 3a and Figure 3c, respectively. For both proteins, peptides appeared equally distributed, no specific hot spot became apparent.

**Figure 3:**
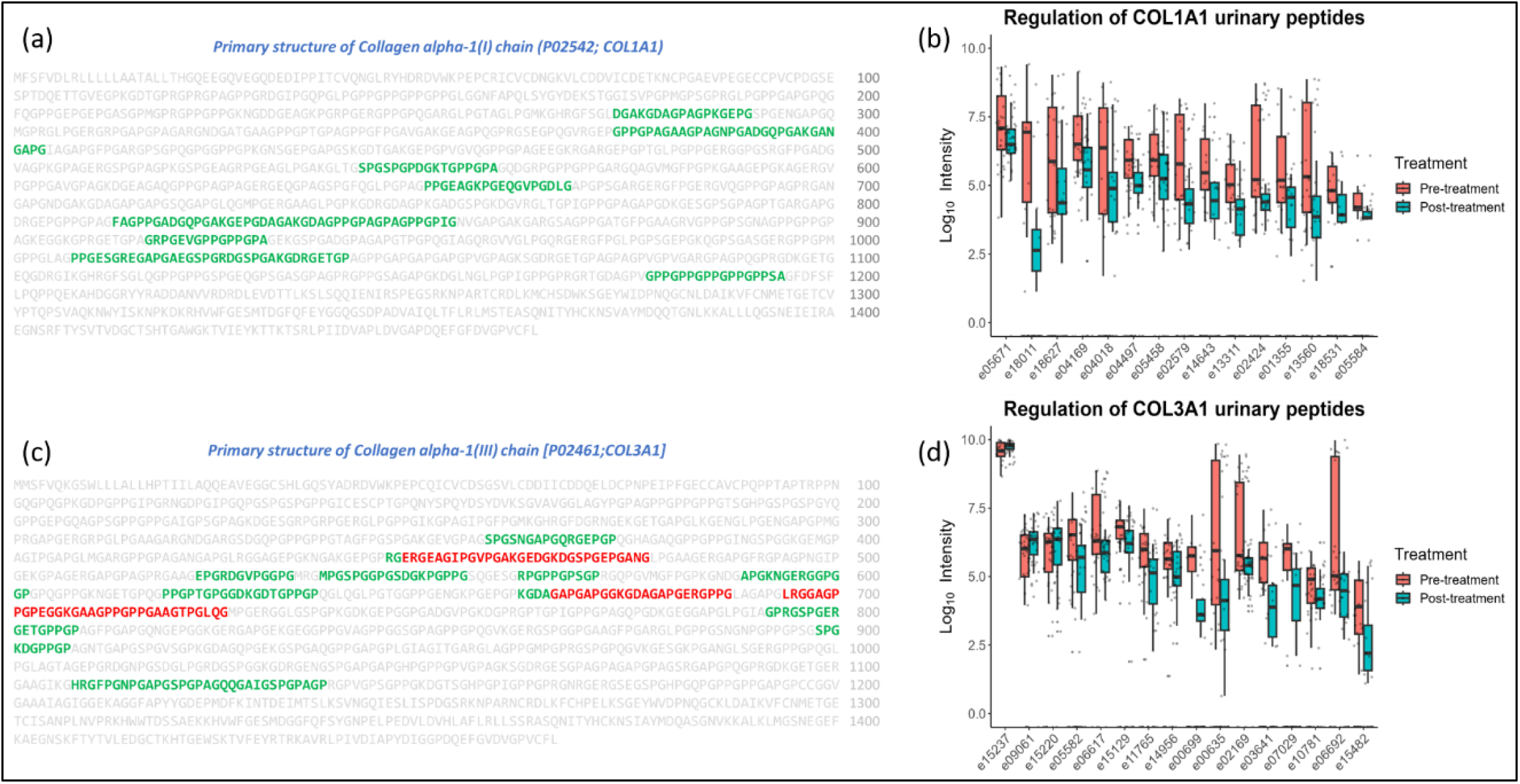
(a) Alignment of identified peptide sequences in this study in the primary structure of protein COL1A1. (b) Box and Whisker plots depicting the down-regulation of all the COL1A1 peptides, in response to GLP-1R agonists treatment. (c) Alignment of identified peptide sequences in this study in the primary structure of protein COL3A1. (d) Box and Whisker plots depicting the up- and down-regulation of all the COL3A1 peptides, in response to GLP-1R agonists treatment. In (a) and (c), the amino acids in green and red depict the down-regulated and up-regulated peptide sequences, respectively.

### Analysis of proteases

To uncover plausible molecular mechanisms responsible for the observed impact of GLP-1R agonists treatment on urinary peptides in T2DM patients, the proteases potentially responsible for cleavage of the 70 statistically significant peptides were investigated using Proteasix. In total, 10 endopeptidases were retrieved as a result of the default search with the “Observed Prediction tool” of Proteasix, putatively responsible for cleaving 38 urinary peptides (36 downregulated and 2 upregulated) out of the 70 urinary peptides. The results are provided in Table S2. Most of the predicted endopeptidases belonged to the matrix metalloproteinase (MMP) family of proteases (7 out of 10 proteases), responsible for cleaving peptides at both the N’ and C’ terminals. Further proteases predicted as potentially responsible for cleaving the N’ terminal belonged to the cathepsin family (CTSL and CTSD) while that cleaving at the C’ terminus was A disintegrin and metalloproteinase with thrombospondin motifs 5 (ADAMTS5).

## Discussion

In the last decade, GLP-1R agonists have been the recommended and preferred second line treatment for T2DM patients. Despite the various advantages of GLP-1R agonists over other anti-hyperglycemic drugs, the underlying molecular mechanisms of treatment with GLP-1R agonists have not been studied in-depth. Aiming to understand the effect of GLP-1R agonist treatment on T2DM patients, the urinary peptidome of thirty-two T2DM patients were analyzed for the first time in this study, using the CE-MS technology. The untargeted peptidomic analysis coupled with statistical tools identified 70 sequenced peptide fragments from 26 proteins, as statistically significantly different (adjusting for multiple testing) in abundance between the pre- and post-treatment samples. Uniform distribution of intensity of the 70 peptides (red spots in Figure 2a), emphasized that their observed significant change with GLP-1R agonist treatment was not a function of abundance in the urine samples. For most of these 70 peptides, a comparative analysis further revealed a combined downregulation with GLP-1R agonist treatment (66/70 peptides).

Along these lines, collectively, all the non-collagen peptides (11 out of 70) showed lower abundance after GLP-1R agonist treatment and each was generated from a different protein namely, SERPINA1, APOC3, CD99, CPSF6, CRNN, SERPINA6, HBA2, MB, VGF, PIGR and TTR. In studies analyzing the effect of Liraglutide (GLP-1R agonist type) on T2DM patients, Rafiullah *et al*.,^36^ reported the downregulation of the urinary protein SERPINA1 and Adiels *et al*.,^37^ reported decreased secretion of APOC3, respectively with treatment. No literature was found reporting regulation of CD99, CPSF6, HBA2, MB and TTR by GLP-1R agonist treatment. However, impact of diabetes and/or obesity on these molecules has been reported: CD99 transcripts have been stated to up-regulate in T2DM profiles^38^; CPSF6 indirectly modulates insulin secretion^39^; HBA2, a commonly known marker for anemia and β-thalassaemia, interferes with glycemic markers of T2DM patients^40^. In addition, elevated levels of MB, a known cardiac marker, were reported in T2DM patients^41^ and increased levels of TTR have been associated with glucose intolerance, obesity and T2DM^42^. Benchoula *et al*.,^43^ in their extensive review reported that VGF is expected to induce obesity, while also playing a role in lipolysis and insulin secretion, hence, acting as a potential target in T2DM therapy. The results in this study may therefore indicate towards the further beneficial effect of GLP-1R agonists in downregulation of CD99, CPSF6, HBA2, MB, TTR and VGF peptides. No evidence on the association of SERPINA6 and PIGR protein with T2DM and/or GLP-1R agonists treatment could be found in the literature.

On the other hand, the remaining 59 out of the 70 statistically significant urinary peptides, majorly generated from three prominent collagen proteins COL3A1 (*n*=16), COL1A1 (*n*=15) and COL1A2 (*n*=10). Recently, He *et al*.,^44^ reported about the high abundance of collagen peptides observed in urine samples, as a result of proline hydroxylation which plausibly inhibits its reabsorption in the kidney. 55 out of the 59 collagen peptides were observed to be significantly downregulated with GLP-1R agonists treatment in the T2DM urinary proteome, while 4 peptides (COL3A1; *n*=3 and COL1A2; *n*=1) showed a significant upregulation on treatment. In-line with the extensive recent report by Mavrogeorgis *et al*.,^45^ all the urinary collagen peptides identified in this study were devoid of the signal peptide, N-terminal pro-peptide and C-terminal pro-peptide; corresponding only to the mature protein region (Figure 3a and 3c). The observed downregulation of the collagen peptides in our study could potentially represent rather an attenuated degradation of the mature collagen protein than resulting from protein synthesis or protein assembling processes.

To further corroborate the above speculation, endopeptidases responsible for putatively cleaving the 70 statistically significant peptides were analyzed with Proteasix. The MMP family of proteases accounted for cleaving the maximum statistically significant peptides (89% of the cleavage sites, i.e., 34 out of the 38 urinary peptides); majorly by MMP2 (21.1%), MMP9 (21.1%) and MMP13 (15.5%). The suggested downregulation in the activity of MMP peptidases as observed by decrease in intensity of collagen peptides in this study, is in-line with literature. Down-regulation in expression of MMP9 was observed on treatment with Liraglutide in a study that included induced-DM rabbit models^46^. In an another study, 45% and 60% reduction in activity of MMP2 and MMP9, respectively, in addition to 60% reduced COL1A1 levels, was observed in a male C57BL/6 mice on treatment with Semaglutide (GLP-1R agonist type)^47^. Research groups exploring the effect of Exenatide (GLP-1R agonist type) treatment on tumor necrosis factor (TNF)-α human coronary artery smooth muscle cells^48^ and human retinal pigment epithelium cells^49^, reported the downregulated expression of MMP2 and MMP9, respectively on treatment. Another study analyzing atherosclerosis associated biomarkers in T2DM female subjects, reported decrease in MMP2 and MMP9 levels, with an increase in GLP-1 and GLP-1R levels^50^. Interestingly, treatment of Human SW1353 with Dulaglutide (GLP-1R agonist type)^51^ and Fibroblast-like synoviocytes cell lines with Exenatide^52^, also resulted in the downregulation of MMP13 and ADAMTS5 proteases.

To conclude, this untargeted peptidomic analysis to identify the effect of GLP-1R agonists treatment on the urinary peptidome of T2DM patients, indicated as a prominent finding the downregulation of MMP proteases, as identified by the downregulation of urinary collagen peptides on GLP-1R agonists treatment. Treatment with GLP-1R agonists also resulted in the decrease of SERPINA1, APOC3, CD99, CPSF6, CRNN, SERPINA6, HBA2, MB, VGF, PIGR and TTR peptides; indicating a potential benefit as many of these proteins express increased levels in T2DM patients. The results also merit the possibility of larger cohort studies to further understand the underlying molecular mechanisms behind the findings.

## Data Availability

All data produced in the present study are available upon reasonable request to the authors.

## Acknowledgments

Author S.L. would like to extend deepest gratitude to the members of Vlahou Lab at BRFAA, Greece and Mosaiques Diagnostics, Germany. Author V.J. is funded by the ‘Deutsche Forschunggemeinschaft’ (DFG, German Research Foundation) through the Transregional Collaborative Research Centre (TRR 219; Project-ID 322900939), (subproject S-03), (INST 948/4S-1); CRU 5011 project number 445703531. Authors H.M., A.V. and V.J. are funded by the Cost-Action CA 21165, IZKF Multiorgan complexity in Friedreich Ataxia, CA201165.

